# The Dawn is Coming —— the Description and Prediction of Omicron SARSCoV-2 Epidemic Outbreak in Shanghai by Mathematical Modeling

**DOI:** 10.1101/2022.04.13.22273788

**Authors:** Minghao Jiang, Guoyu Meng, Geng Wu

**Affiliations:** Shanghai Institute of Hematology, State Key Laboratory of Medical Genomics, National Research Center for Translational Medicine, Rui-Jin Hospital, Shanghai Jiao Tong University School of Medicine and School of Life Sciences and Biotechnology, Shanghai Jiao Tong University, 197 Ruijin Er Road, Shanghai 200025, China; State Key Laboratory of Microbial Metabolism, School of Life Science and Biotechnology, The Joint International Research Laboratory of Metabolic & Developmental Sciences, Shanghai Jiao Tong University, Shanghai, China

**Keywords:** SARS-CoV-2 Omicron, Shanghai outbreak, Prediction model

## Abstract

The COVID-19 Omicron outbreak in Shanghai has been going on for >1 month and 25 million population is subjected to strict lock-down quarantine. Until now, it is not clear how long this epidemic might end. Here, we present a time-delayed differentiation equation model to evaluate and forecast the spreading trend. Our model provides important parameters such as the average quarantine ratio, the detection interval from being infected to being tested positive, and the spreading coefficient to better understand the omicron progression. After data fitting, we concluded on 11 April that the maximum overall number infected in Shanghai would exceed 300,000 on 14 April and the turning point would be in the coming days around 13-15 April, 2022, which is perfectly in line with the real-life infection number. Furthermore, the quarantine ratio in Shanghai was found to be greater than 1, supportive of the effectiveness of the strict lockdown policy. Altogether, our mathematical model helps to define how COVID-19 epidemic progresses under the Shanghai lock-down unprecedented in human history and the Chinese zero tolerance policy.

## Introduction

The SARS-CoV-2 Omicron variant, reported as a new variant of COVID-19 in November, 2021^1^, has broken out in Shanghai recently. This variant carries mutations to help the virus to resist or escape immunity provided by COVID-19 vaccines ^1-3^.To prevent the spread of infection, the local government of Shanghai municipal decided to enforce a practical lockdown on 28 March. However, in the past one and a half weeks, there were still over 20,000 Omicron cases confirmed per day, including both diagnosed and asymptomatic. The situation was quite severe when compared to other places across mainland China during the same period or that in Shanghai several months ago. Here, we propose a new mathematical model to evaluate the epidemic trend of Omicron spreading in Shanghai and make forecasts for the near future.

## Results

The Omicron epidemic in Shanghai has begun since early March, 2022. Until 12 March, the number of confirmed cases per day did not exceed 100 (**Extended Data Fig. 1**). Due to the surge in COVID-19 positives since mid-March, the Shanghai municipal government made the practical lockdown decision on March 28 to control the spread.

We obtained a time-delayed differential equation to describe the spreading of COVID-19 Omicron (**Fig. 1**):

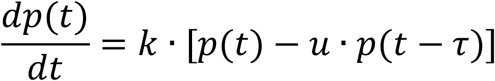

In the above equation, *p*(*t*) is the total number of patients infected at time*t*. The average spreading coefficient *k* represents the average number of people one unquarantined patient can infect in a unit interval. The average quarantine ratio *u* is defined as the number of quarantined patients divided by the number of people showing nucleic acid positive at time *t*. The average detection interval *τ* is the average time interval from being infected to being tested positive.

**Figure 1.**
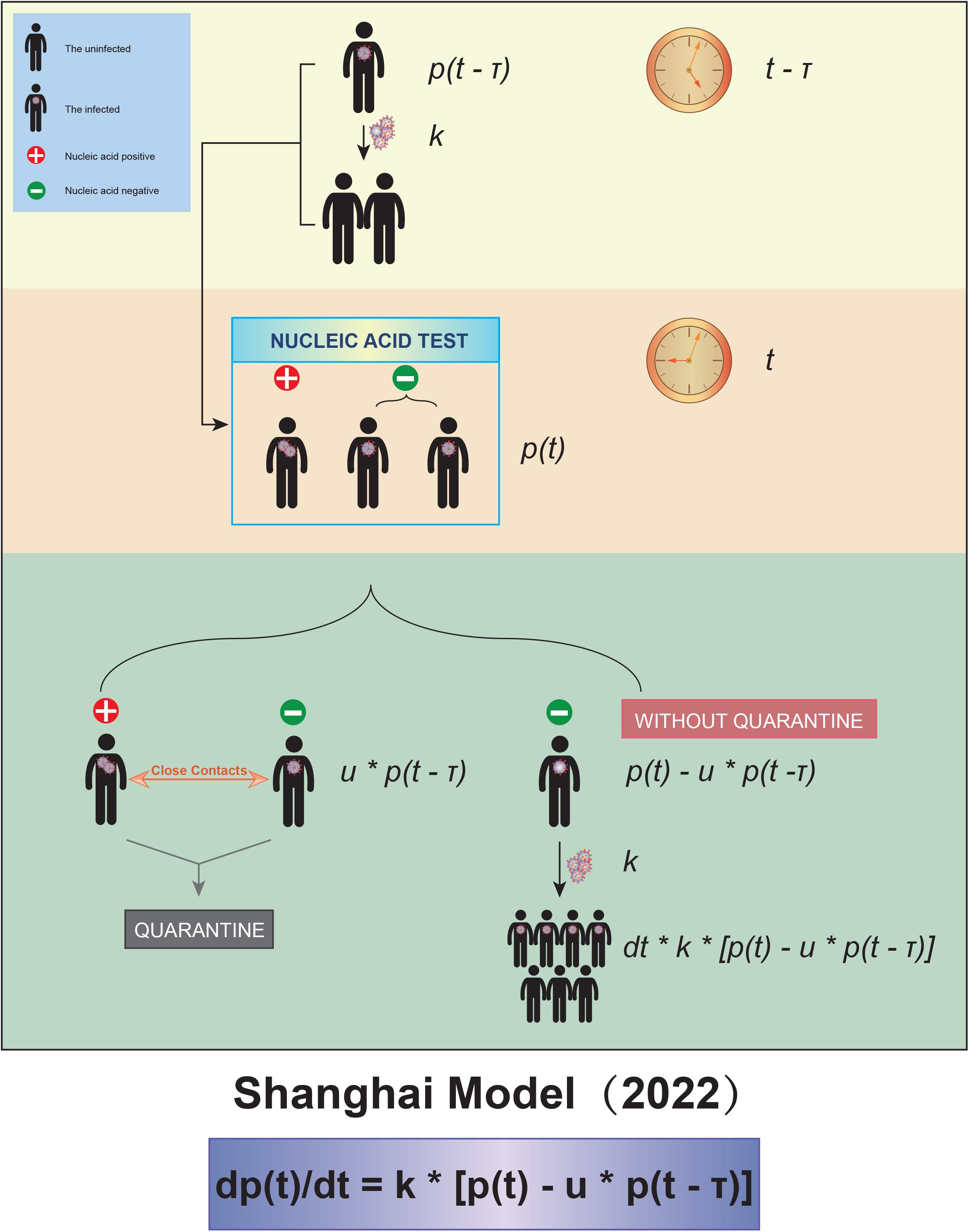
The schematic diagram of our model. Three parameters were taken into account, the test interval *τ*, the spreading coefficient *k* and the quarantine ratio *u*.

We fitted the data of total number of reported cases before and after 28 March to our equation above, in order to figure out to what extent the more stringent policy could affect. We found that the fitting between the reported data and the curve calculated from our equation was quite close (**Fig. 2**). We found that, in order to fit the data better, it was needed to alter the parameter *τ* in the two scenarios: i) *τ* = 2 (days) before the city lockdown and ii) *τ* = 1 (day) after the city lockdown. This can be construed as that the more stringent quarantine policy after the city lockdown, with nucleic acid tests for all citizens once or twice per day, reduced the detection interval *τ*.

**Figure 2.**
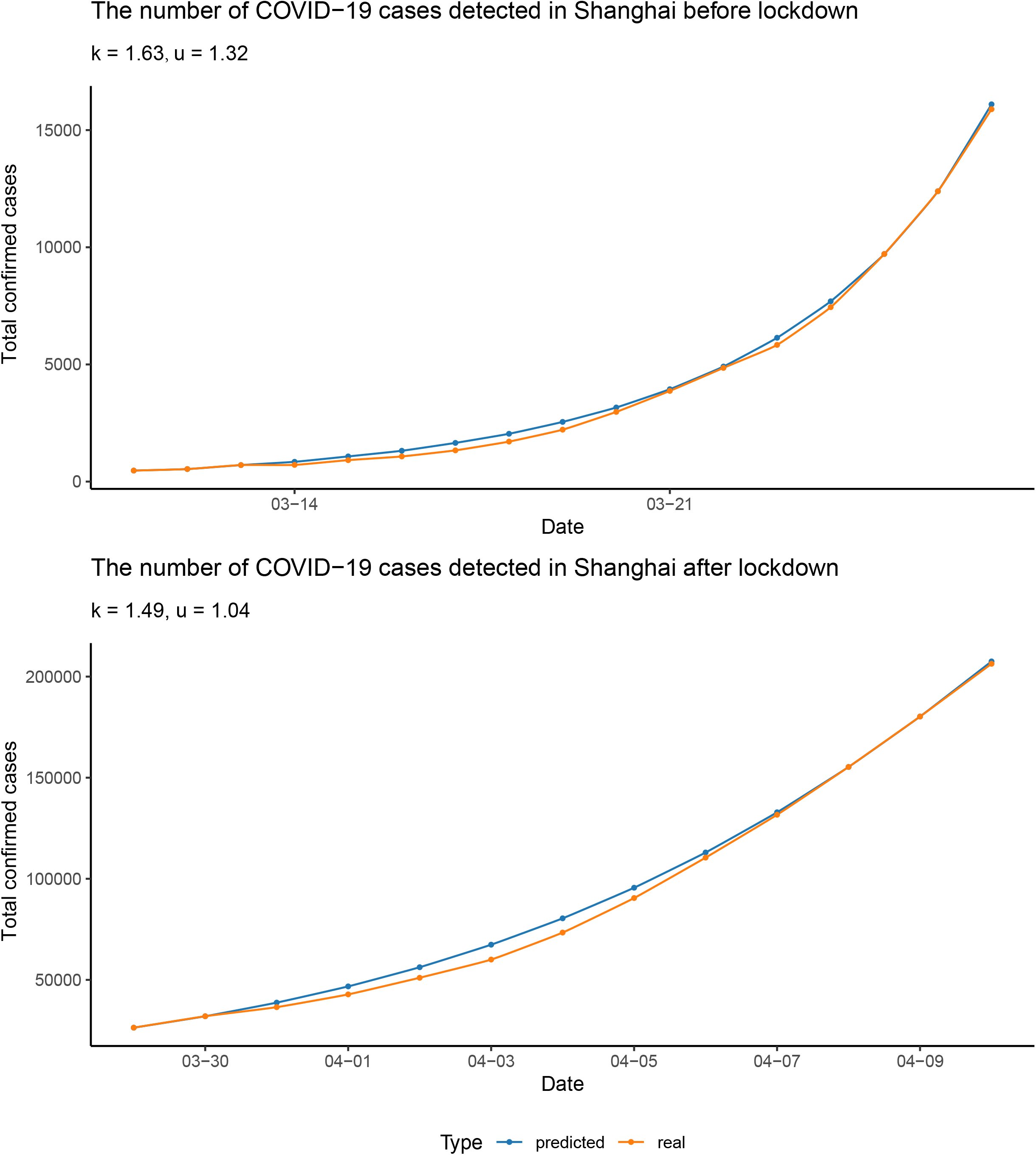
Data fitting. The data were fitted separately before and after the lockdown. The blue points and curve represented the reported number of total confirmed cases, and the yellow ones represented the number calculated from our differential equation. The parameters were rounded to 3 significant digits.

In addition, we found that the spreading coefficient *k* decreased after the city lockdown, from 1.63 to 1.49, approximately (**Fig. 2**). Furthermore, in the early stage of the epidemic, the quarantine ratio *u* was 1.32 > 1, which means that almost all the patients tested positive as well as their close contacts were quarantined. In the later stage of the epidemic, the quarantine ratio *u* became smaller, ∼1.04. The reduced *u* may serve as a sign of the arrival of the turning point. Since *p*(*t*), the cumulative number of confirmed cases, is a non-decreasing function, its derivative,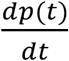, has to be greater than or equal to 0. Therefore, from our equation, *u* must be smaller than or equal to 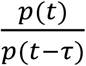. At the beginning of the epidemic, Omicron was spreading quickly,*p*(*t*) was much larger than *p*(*t* − *τ*), so *u* was relatively bigger. When the turning point was getting close, Omicron spreading was slowed down, thus *p*(*t*) was not much different from 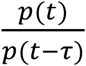 was close to 1, which made the quarantine ratio *u* smaller than that in the early stage of the epidemic.

With the three parameters estimated, the trend of Omicron spreading in the near future was predicted. We concluded that there will be more than 300,000 people infected in Shanghai during this Omicron outbreak (**Fig. 3a**). Most importantly, we predicted that the number of confirmed cases per day would start to decrease around April 13 (**Fig. 3b**), which was the so-called turning point.

**Figure 3.**
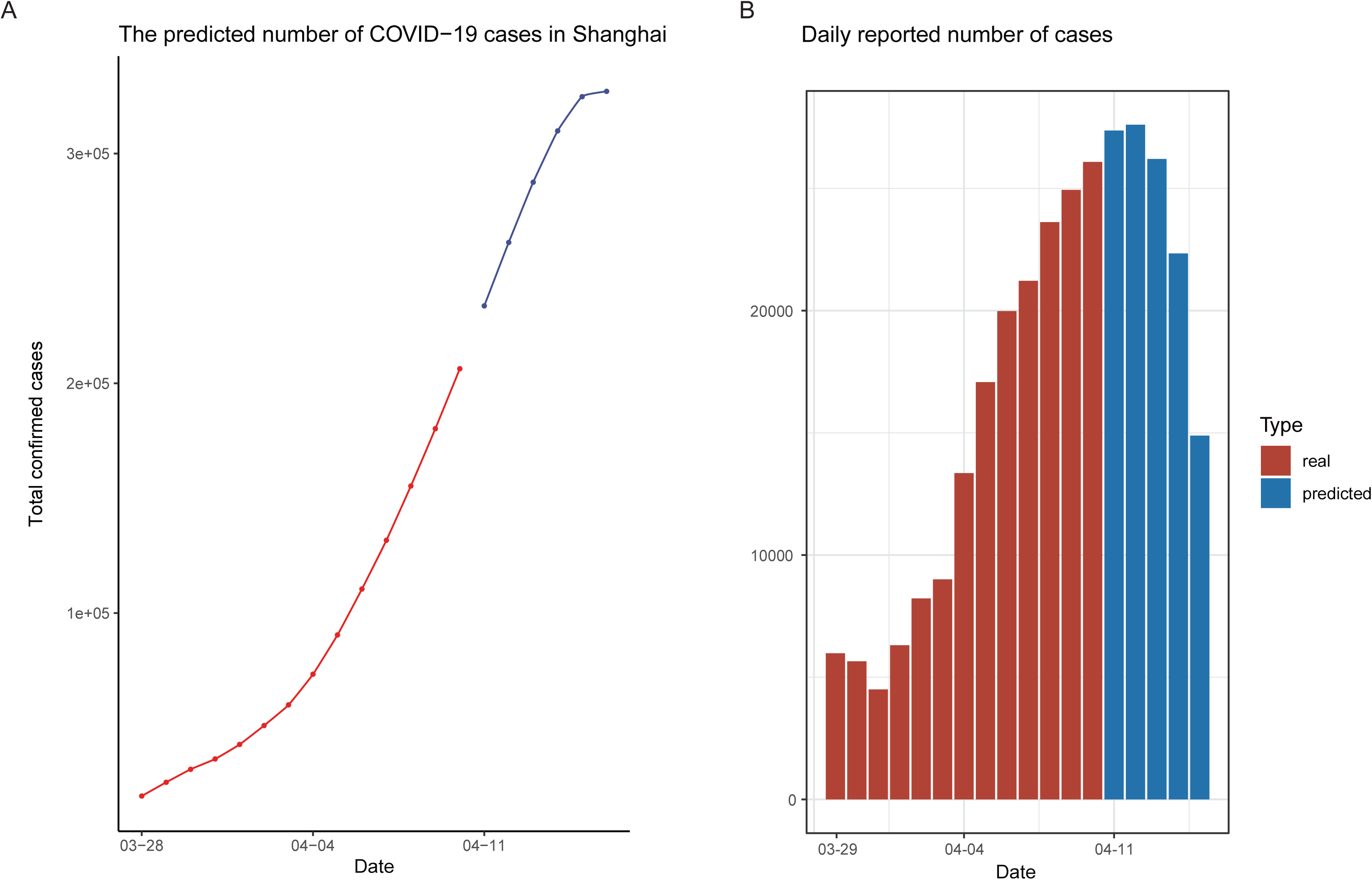
Epidemic trend forecast. **a**, The forecasted total number of confirmed cases, with the predicted data points colored in blue. **b**, The reported daily increased number of cases (red) and those for upcoming days predicted by our model (blue).

We found that the average detection interval *τ* was 1 or 2 days when fitting the known data, which means that, there would be 1 to 2 days of delay from infection to showing positive in the nucleic acid test, during which time the unnoticed patients were still infectious^4-6^. We believe that this is one of the major reasons why Omicron spread rapidly. Furthermore, the average quarantine ratio *u* was found to be greater than 1, which means that most of the infected patients and their close contacts had been quarantined, indicating that the lockdown and other stringent policies were effective, making appearance of the turning point and quelling the epidemic possible.

Moreover, the apparent R0 value, which is the number of people a patient could infect, could be obtained by multiplying the spreading coefficient *k* and the detection interval *τ*. After the lockdown, the value of R0 dropped from 3.27 to 1.49, providing further supporting evidence for the effectiveness of the lockdown.

We finished the forecast on April 10. Before our manuscript is submitted, the numbers of daily infected patients before April 17 were collected to check our prediction (**Supplementary Table 2**). Strikingly, it turned out that the real turning point indeed appeared on 13 April, showing exact concordance with our prediction.

## Discussion

The sudden Omicron epidemic outbreak in Shanghai has brought panic to the public, and great loss to business. In order to evaluate and forecast the spreading trend based on available confirmed data, we proposed a novel mathematical model, which took the Chinese government policy (such as frequent nucleic acid tests and isolating those showing nucleic acid positive as well as their close contacts) into account, which could provide us important parameters describing the Omicron epidemic such as the spreading coefficient *k*, the quarantine ratio *u*, and the detectioninterval *τ*. The predicted number of overall cases and the expected time of turning point may help the government to make judgment on the spreading and to revise the policies accordingly.

Although our mathematical model is forged based on China’s anti-epidemic strategies which rely heavily on quarantine, it is also applicable to other countries (including western countries) in the world. Compared to western countries relying on the formation of herd immunity and mRNA vaccines which reduce the average spreading coefficient *k*, China attempts to boost the average quarantine ratio *u* to get the epidemic under control.

Our model is easy to be applied to describe the spreading of other epidemic diseases, and can help the society to avoid panic and build up confidence to fight against COVID-19. For example, the Guangzhou city was the latest metropolitan city in China that declared city-wise lockdown on Apr 11. We believe that our mathematical model presented in this report would help the public to have a better grasp of the current pandemic spread, and would undoubtedly instill confidence and calm that are urgently needed in the caught-up fight of COVID-19 in China.

## Materials and methods

### Data used in modeling

The case numbers were collected manually from officially released reports (**Supplementary Table 1**).

### Mathematical modeling

First, a differential equation was deducted to describe the spreading of COVID-19 as following. Let us denote *s*(*t*) as the number of people whose nucleic acid tests are positive, then

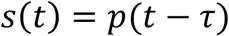

in which *p*(*t*) is the total number of patients infected at time *t*, and *τ* is the time interval from being infected to being tested positive. Since the patients infected during the time *τ* will not be recorded as nucleic acid positive, the diagnosed or recorded patients (*s*(*t*) above) at time *t* is equal to the number of infected population at time *t* − *τ*.

During the time *dt*, the number of newly infected patients *dp*(*t*) is

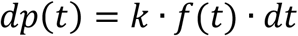

Where *k* is the spreading coefficient representing the average number of people one unquarantined patient can infect in a unit interval, and *f*(*t*) is the number of unquarantined patients at time *t*. *f*(*t*) is equal to *p*(*t*) subtracted by the number of quarantined patients*q*(*t*):

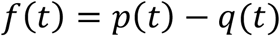

We further assume that *q*(*t*) (including both nucleic acid positive patients and their close contacts) is proportional to *s*(*t*), and let *u* be the average quarantine ratio:

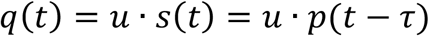

Therefore, we get the final differential equation:

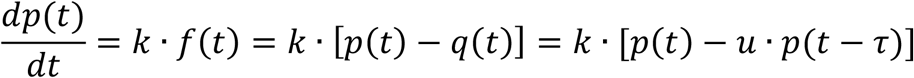

In this equation, there are three unknown parameters: the average spreading coefficient *k*, the average quarantine ratio *u*, and the average detection interval *τ*. These parameters may vary during different periods, *e*.*g*., before or after the city lockdown. Using the number of reported cases during a particular time in Shanghai, these three parameters can be estimated by data fitting. As a next step, we could forecast the trend of Omicron spreading in Shanghai with these estimated parameters.

### Additional information

This manuscript/model was submitted to on-line preprint server (https://www.medrxiv.org) on 12 Apr (using infection data available up to 11 Apr). Now, on the eve of our latest submission to Nature on 19 Apr, we can check the real-life infection against our prediction. As shown in **Supplementary Table 2** and **Fig. 4**, the actual turning point is on 13 Apr and the date when the total infection number exceeded 300,000 is on 14 Apr, perfect matches to our mathematical description/prediction.

**Figure 4.**
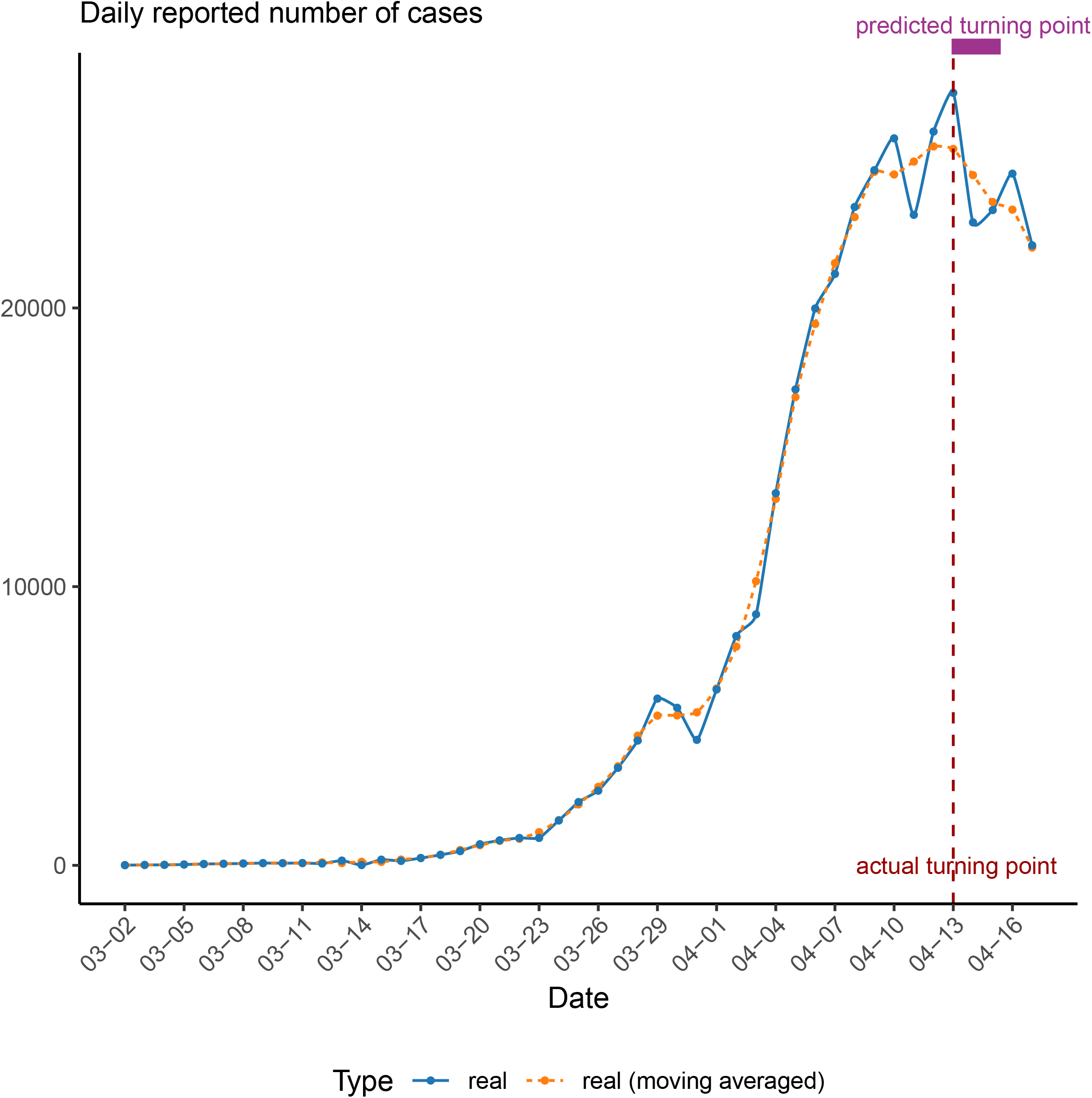
The number of infected cases per day. The dashed line represented the moving averaged number of real cases while the solid line represented the number of real cases. The actual and predicted turning point were represented by the vertical dashed line and the rectangle above the curves.

## Data Availability

All data produced in the present study are available upon reasonable request to the authors

## Acknowledgements

This work was supported by research grants 81970132, 81770142, 81800144, 31800642, 82100153 and 82104582 from National Natural Science Foundation of China, a research grant 20JC1410600 from Shanghai Science and Technology Committee, Shanghai Guangci Translational Medical Research Development Foundation, a research grant 20152504 from “Shanghai Municipal Education Commission—Gaofeng Clinical Medicine Grant Support”, “The Program for Professor of Special Appointment (Eastern Scholar) at Shanghai Institute of Higher Learning”, Samuel Waxman Cancer Research Foundation. We thank Professor Zhu Chen, Shanghai Institute of Hematology, for his kind support of this work.

## Author contributions

Conceived and designed the experiments: GM and GW. Performed the experiments: MJ. Analyzed the data, preparation of figures manuscripts and wrote the paper: MJ, GM, GW. Project supervision: GM. All authors read and approved the final manuscript.

## Compliance with ethics guidelines Conflict of interest

This article does not contain any studies with human or animal subjects. The authors declare no competing interests.

## Extended Data

**Figure 1.**
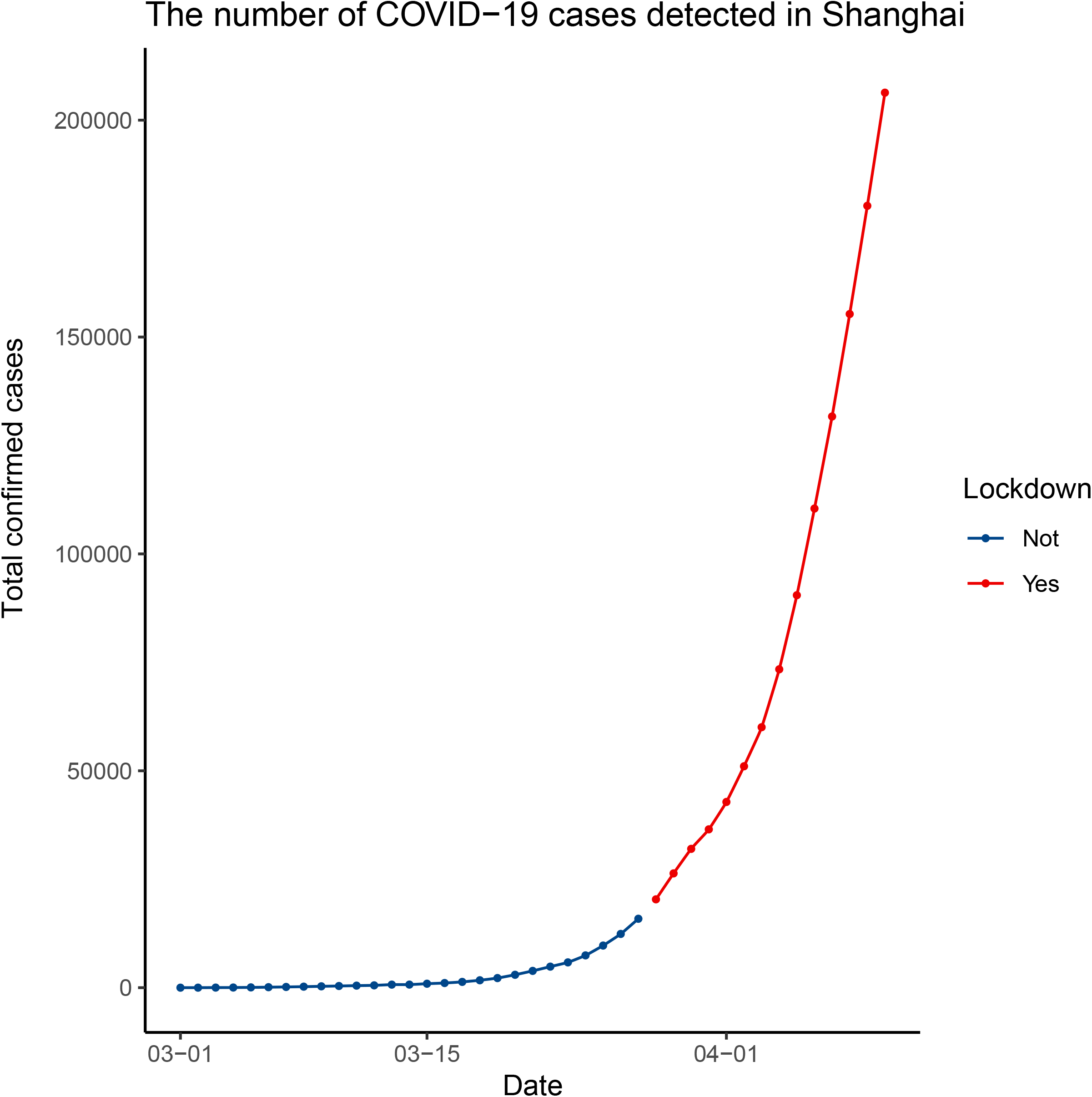
The total number of confirmed cases in Shanghai since March1. The blue points and curve represent the data before the official lockdown, and the red ones represent the data after the lockdown.

**Supplementary Table 1.** The number of predicted and real infected cases.

| predicted | real total infected | real daily reported | real total infected (*) | real daily reported (*) | date |
| --- | --- | --- | --- | --- | --- |
| 233705.4 | 229678 |  | 230672 | 25253 | 4/11 |
| 261313.8 | 256008 | 26330 | 256469 | 25797 | 4/12 |
| 287521 | 283727 | 27719 | 282176 | 25707 | 4/13 |
| 309862.9 | 306799 | 23072 | 306944 | 24768 | 4/14 |
| 324750.5 | 330312 | 23513 | 330745 | 23801 | 4/15 |
| 327074 | 355132 | 24820 | 354272 | 23527 | 4/16 |
|  | 377380 | 22248 | 376442 | 22170 | 4/17 |

\*These numbers were moving averaged

**Supplementary Table 2.** The number of COVID-19 cases in Shanghai.

| **Daily infected (\*)** | **Total infected (\*)** | **Date** |
| --- | --- | --- |
| 1 | 1 | 3/1 |
| 5 | 6 | 3/2 |
| 14 | 20 | 3/3 |
| 16 | 36 | 3/4 |
| 28 | 64 | 3/5 |
| 48 | 112 | 3/6 |
| 55 | 167 | 3/7 |
| 65 | 232 | 3/8 |
| 80 | 312 | 3/9 |
| 75 | 387 | 3/10 |
| 83 | 470 | 3/11 |
| 65 | 535 | 3/12 |
| 169 | 704 | 3/13 |
| 9 | 713 | 3/14 |
| 202 | 915 | 3/15 |
| 158 | 1073 | 3/16 |
| 260 | 1333 | 3/17 |
| 374 | 1707 | 3/18 |
| 509 | 2216 | 3/19 |
| 758 | 2974 | 3/20 |
| 896 | 3870 | 3/21 |
| 981 | 4851 | 3/22 |
| 983 | 5834 | 3/23 |
| 1609 | 7443 | 3/24 |
| 2269 | 9712 | 3/25 |
| 2678 | 12390 | 3/26 |
| 3500 | 15890 | 3/27 |
| 4477 | 20367 | 3/28 |
| 5982 | 26349 | 3/29 |
| 5653 | 32002 | 3/30 |
| 4502 | 36504 | 3/31 |
| 6311 | 42815 | 4/1 |
| 8226 | 51041 | 4/2 |
| 9006 | 60047 | 4/3 |
| 13354 | 73401 | 4/4 |
| 17077 | 90478 | 4/5 |
| 19982 | 110460 | 4/6 |
| 21222 | 131682 | 4/7 |
| 23624 | 155306 | 4/8 |
| 24943 | 180249 | 4/9 |
| 26087 | 206336 | 4/10 |

\* Data source: Shanghai Municipal Health Commission (https://wsjkw.sh.gov.cn/xwfb/index.html) and Global Times (https://www.huanqiu.com)

